# “A longitudinal study of healthcare workers’ surveillance during the ongoing COVID-19 Epidemics in Italy: is SARS-CoV-2 still a threat for the Health-care System?”

**DOI:** 10.1101/2021.02.23.21249481

**Authors:** F Barbaro, F Della Rocca, A Padoan, A Aita, V Cianci, D Basso, A Cattelan, D Donato, M Plebani, L Dall’Olmo

## Abstract

**Objectives:** In spring 2020, Northern Italy was the first area outside China to be involved in the SARS-CoV-2 pandemic. This observational study depicts SARS-CoV-2 prevalence and serological curves among *first-line* healthcare workers (HCWs) at Padua University Hospital (PdUH), North-East Italy.

**Method:** 344 HCWs, working at the PdUH Emergency Department and Infectious Disease Unit, underwent a SARS-CoV-2 RNA nasopharyngeal swab with paired IgM and IgG antibody detection for 4 consecutive weeks. At every session, a questionnaire recorded symptoms, signs and recent contacts with SARS-CoV-2 patients. Positive cases were followed up for 5 months.

**Results:** Twenty-seven HCWs (7.84%) had positive serology (Abs) with 12 positive swabs during the study period. Two additional HCWs were positive by swab but without Abs. Fourteen cases (4%) had SARS-CoV-2 infection before the beginning of the study. An HCW with autoimmune disease showed false Ab results. 46% of individuals with Abs reported no symptoms, in accordance with previous population studies. Fever, nasal congestion, diarrhoea and contacts with SARS-CoV-2 individuals correlated to SARS-CoV-2 infection. 96% of Abs+ cases showed persistent positive antibodies 5 months later and none was re-infected.

**Discussion:** Correct use of PPEs and separate paths for positive/negative patients in the hospital can result in a low percentage of SARS-CoV-2 infections among HCWs, even in high risk settings. Frequent testing for SARS-CoV-2 with nasopharyngeal swabs is worthwhile, irrespective of HCWs’ symptoms, due to the lack of specificity together with the high percentage of asymptomatic cases. Further studies are needed to elucidate the neutralizing effect of SARS-CoV-2 antibodies.

## INTRODUCTION

### Background/Rationale

The new Coronavirus disease 2019 (COVID-19) pandemic represents the most serious global challenge after the Second World War, with many publichealth and economic consequences. The SARS-CoV-2 epidemic spread out in December 2019 in the Wuhan Province of Hubei, China, and Italy was the first country outside Asia to be involved. The municipality of Vo’ Euganeo, 30 Km from PdUH, was recognized as the first COVID-19 cluster in Europe after the first confirmed death, the 27^th^ of February 2020. More than 200 Italian physicians have died of COVID-19 up to the end of November 2020. While waiting for an available and efficient vaccine against the virus, prevention of interpersonal diffusion remains the most effective measure to limit viral spread. SARS-CoV-2 transmission among HCWs remains a significant concern to date, not only for the risk of HCWs of becoming infected but also infecting patients, co-workers and family members. It should also be taken into careful consideration that transmission might occur from symptomatic, but mainly from asymptomatic subjects. SARS-CoV-2 diagnosis should be confirmed by positive molecular results of nasopharyngeal swabs, while serology remains a relevant laboratory test to identify previous or silent infections.

### Objectives

During the COVID-19 outbreak in spring 2020, this longitudinal study followed up HCWs of 3 *first-line* wards of PdUH,in order to estimate: 1) the incidence of SARS-CoV-2 infections and/or illnesses among HCWs by rRT-PCR from nasopharyngeal swabs; 2) the *time-course* of SARS-CoV-2 IgM and IgG serological determinations.

## MATERIALS AND METHODS

### Study design

This prospective study was carried out between April 8 and May 29, 2020 (week 15–22 of 2020) at PdUH, a tertiary-care hospital in the Veneto Region (Northeast Italy). In February 2020, PdUH developed an emergency plan for the COVID-19 outbreak that included a COVID-19 triage prior to entry to the hospital and dedicated areas for COVID-19 suspected and confirmed cases (1). HCWs were provided with personal protective equipment (PPE) according with eCDC and National Health recommendations (2, 3) and trained for their proper use. A hospital surveillance protocol was instituted to monitor infections among HCWs.

The study settings were:

a. Padua University Hospital Emergency Unit, Pronto Soccorso Azienda Ospedale Università Padova(PS AOUP)
b. Padua University Hospital ancillary Emergency Unit at Saint Anthony City Hospital, Pronto Soccorso Ospedale Sant’Antonio, Padova (PS OSA)
c. Padua University Hospital Infectious Diseases Unit (IDU), composed of ward and Advanced Triage (AT).

The IDU ward was dedicated to the admission of confirmed COVID-19 patients, while the IDU AT was a temporary outpatient clinic for the first evaluation of mildly symptomatic patients and the execution of nasopharyngeal swabs in tents. (1).

A total of 344 HCWs underwent SARS-CoV-2 RNA nasopharyngeal swabs with paired IgM and IgG antibody detection, once a week for 4 consecutive weeks. At every session, a questionnaire was administered to participants, recording the demographic characteristics, the professional role, the presence of comorbidities and clinical symptoms and signs (e.g., fever, cough, nasal congestion, sore throat, diarrhoea, dyspnea) and eventual contacts with SARS-CoV-2 patients in the last week. According to nasopharyngeal swab and SARS-CoV2 serological test results, participants were divided in four classes:

– Positive cases to swab only (class 1)
– Positive cases to serology only (class 2)
– Double positive cases (class 3)
– Double negative participants (class 4)

Only positive participants for SARS-CoV-2 serology were followed up for 5 months and tested at the end of this period with both nasopharyngeal swabs and serology. All participants were informed about the purpose and procedures of the study and gave informed consent. The study was conducted in accordance with the Declaration of Helsinki. Participation was voluntary; subjects could withdraw at any time and all analyses were carried out on anonymized data. HCWs were evaluated for body temperature and symptoms at the beginning of every shift.

### Diagnostic tests

#### Nasopharyngeal swabs

nasopharyngeal swabs were performed by using flocked swabs in a liquid-based collection and transport system (eSwab®, Copan Italia Spa, Brescia, Italy). All nasopharyngeal swab samples were processed with an in-house Real-Time Polymerase Chain Reaction (RT-PCR) method according to Lavezzo et al. (4). All tests were performed at the Clinical Microbiology and Virology Unit of PdUH, which is the regional reference laboratory for emerging viral infection.

#### Serologic test

SARS-CoV-2 IgM and IgG immune response was evaluated according to Padoan et al. (5) with the MAGLUMI™ 2000 Plus method. MAGLUMI™ 2000 Plus (New Industries Biomedical Engineering Co., Ltd [Snibe], Shenzhen, China) is a chemiluminescent analytical system (CLIA) featuring high throughput (up to 180 tests/h). According to the manufacturer’s instructions (271 2019-nCoV IgM, V2.0, 2020-03 and 272 2019-nCoV IgG, V1.2, 2020-02), the 2019-nCoV IgM cut-off is 1.0 AU/mL, while the 2019-nCoV IgG cut-off is 1.1 AU/mL. The manufacturers claim that the calculated clinical sensitivities of IgM and IgG are 78.65% and 91.21%, respectively, while specificities of IgM and IgG are 97.50% and 97.3%, respectively. All serological tests were performed at the Laboratory Medicine Unit of PdUH.

### Statistical Analysis

Statistical analyses were performed using Stata v16.1 (Statacorp, LakeWay drive, TX, USA). Mean and standard deviation or median and interquartile range and percentages were used as descriptive statistics for normally distributed, or skewed distributed variables, as appropriate. Fisher’s exact test was employed to evaluate differences across groups in categorical variables. To assess differences among groups, T-test and ANOVA were performed with continuous data. Exact logistic regression was employed to define the association between studied variables and SARS-CoV-2 positive testing. The user community command ‘xtgraph’ was used to plot time kinetics of IgM and IgG antibodies in the study period, while spaghetti plots were plotted using Stata native command ‘xtline’.

## RESULTS

### Study population characteristics

A total of 344 HCWs were enrolled in the study; the percentage of HCW participation was high (95,3%; 344/361). One hundred twenty-six individuals (36.6%) were accrued from PS AOUP, 52 (15.1%) from PS OSA and 166 (48.3%) from IDU. Among the 344 subjects enrolled in the study, 339/344 (98.5%) individuals were followed up for 1 week, 327/344 (95.1%) for 2 weeks, 312/344 (90.7%) for 3 weeks, respectively. A series of 25 subjects positive to IgG and/or IgM test were re-tested 5 months later for serum antibodies. HCWs’ professional qualifications were different across wards (Fisher’s exact test = 0.002), varying for clinicians from 21.2% PS OSA to 23.5% IDU to 38.9% PS AOUP, for nurses from 50% PS OSA to 50.8%PS AOUP to 53% IDU, for healthcare assistants from 10.32% PS AOUP to 23.5% IDU to 28.8% PS OSA. Gender did not differ across wards considering clinicians (Fisher’s exact test, p = 0.649), considering nurses (Fisher’s exact test, p = 0.144) or considering healthcare assistants (Fisher’s exact test, p = 0.748). Differently, gender differed across professional qualifications (Fisher’s exact test, p < 0.001), being the overall number (and percentages) of females 54/99 (54.5%) for clinicians, 127/177 (71.7%) for nurses and 55/67 (82.1%) for healthcare assistants. Fig 1 shows age differences by gender and wards, for clinicians, nurses and healthcare assistants. From analysis of variance (ANOVA), considering gender, wards and professional qualifications as covariates, age was associated with professional qualification (F = 10.83, p < 0.001), clinicians being younger than nurses or healthcare assistants.

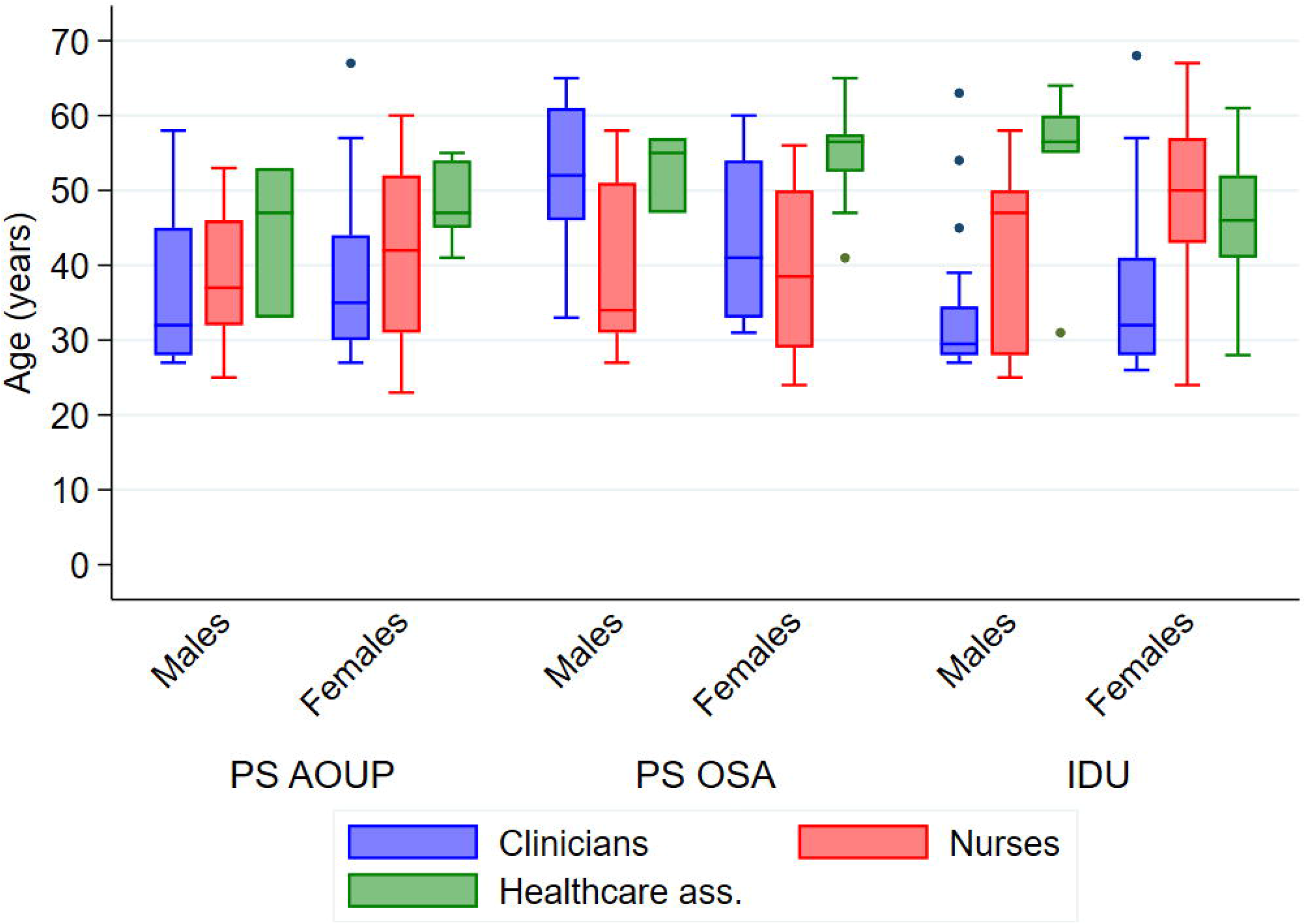

### Evaluation of the association between SARS-CoV-2 and the studied variables

A total of 14/344 (4.07%) were positive to SARS-CoV-2 nasopharyngeal swab testing throughout the study period.SARS-CoV-2 diagnosis was not associated with wards (Fisher’s exact test, p = 0.106), with professional qualifications (Fisher’s exact test, p = 0.089) or with gender (Fisher’s exact test, p = 0.071). Tab 1 reports the association of SARS-CoV-2 positivity with the other studied variables (age, symptoms and previous diseases) and with serological results.

**Tab 1:**
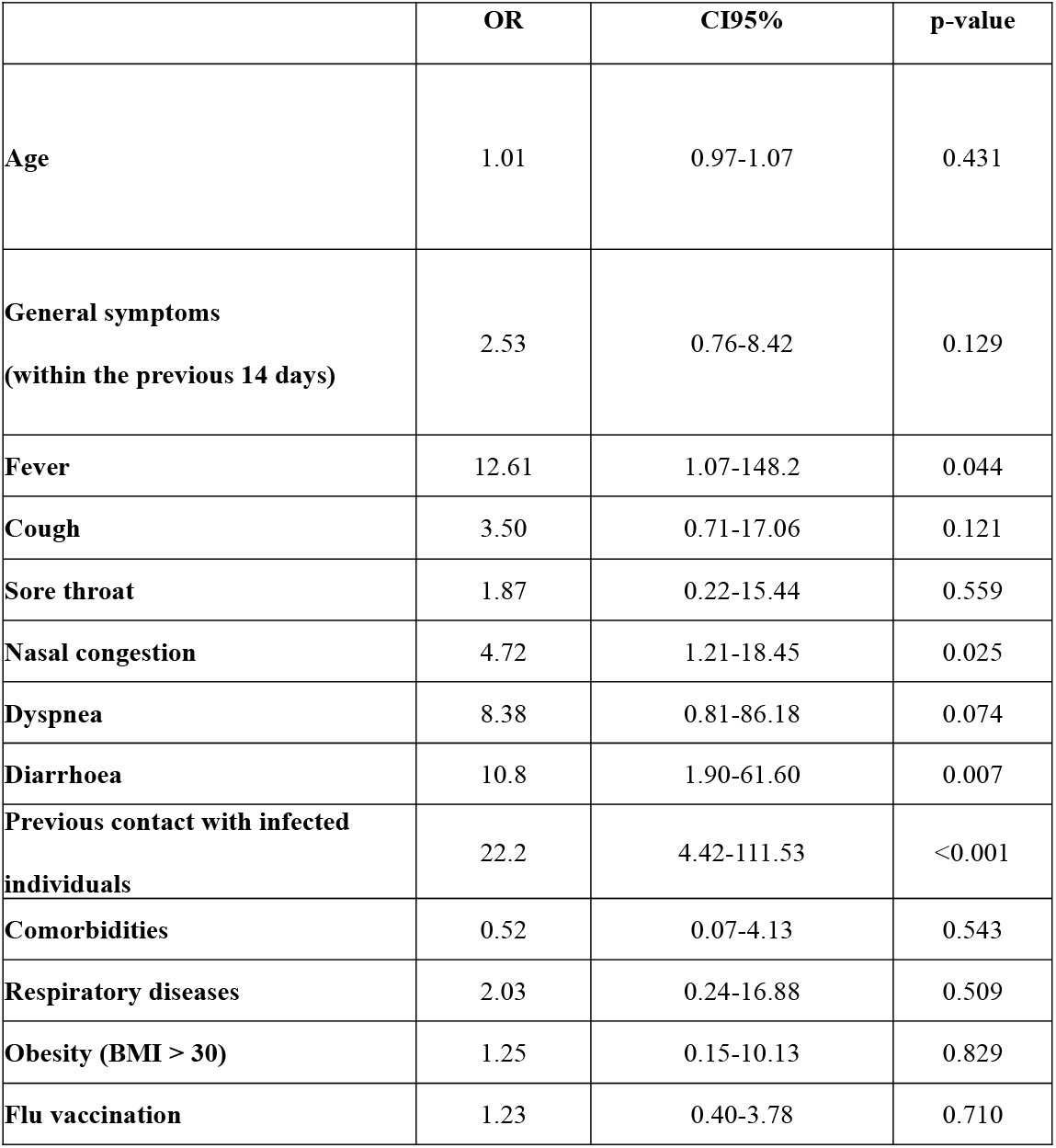
Exact logistic regression results [Odds Ratios (OR) and CI95%] of the studied variables with SARS-CoV-2 positivity

|  | **OR** | **CI95%** | **p-value** |
| --- | --- | --- | --- |
| **Age** | 1.01 | 0.97-1.07 | 0.431 |
| **General symptoms**<br>**(within the previous 14 days)** | 2.53 | 0.76-8.42 | 0.129 |
| **Fever** | 12.61 | 1.07-148.2 | 0.044 |
| **Cough** | 3.50 | 0.71-17.06 | 0.121 |
| **Sore throat** | 1.87 | 0.22-15.44 | 0.559 |
| **Nasal congestion** | 4.72 | 1.21-18.45 | 0.025 |
| **Dyspnea** | 8.38 | 0.81-86.18 | 0.074 |
| **Diarrhoea** | 10.8 | 1.90-61.60 | 0.007 |
| **Previous contact with infected**<br>**individuals** | 22.2 | 4.42-111.53 | <0.001 |
| **Comorbidities** | 0.52 | 0.07-4.13 | 0.543 |
| **Respiratory diseases** | 2.03 | 0.24-16.88 | 0.509 |
| **Obesity (BMI > 30)** | 1.25 | 0.15-10.13 | 0.829 |
| **Flu vaccination** | 1.23 | 0.40-3.78 | 0.710 |

At the beginning of the study (t0), the number (and percentages) of individuals with positive IgM was 6 (1.74%), with positive IgG was 25 (7.27%) and with positive IgM or IgG was 27 (7.85%). At t0, IgG and/or IgM antibodies were positive in 12/14 (85.7%) SARS-CoV-2 positive subjects and in 15/330 (4.55%) negative subjects. Fig 2A shows the variations of individual IgG values measured in the 14 SARS-CoV-2 positive subjects. Fig 2B shows the time course kinetics of IgM and IgG in SARS-CoV-2 positive and negative subjects. Antibody-positive HCWs were invited to repeat testing after the end of the study at five months. Twenty-five subjects responded positively, and among them, 24 had persistent positive antibody results.

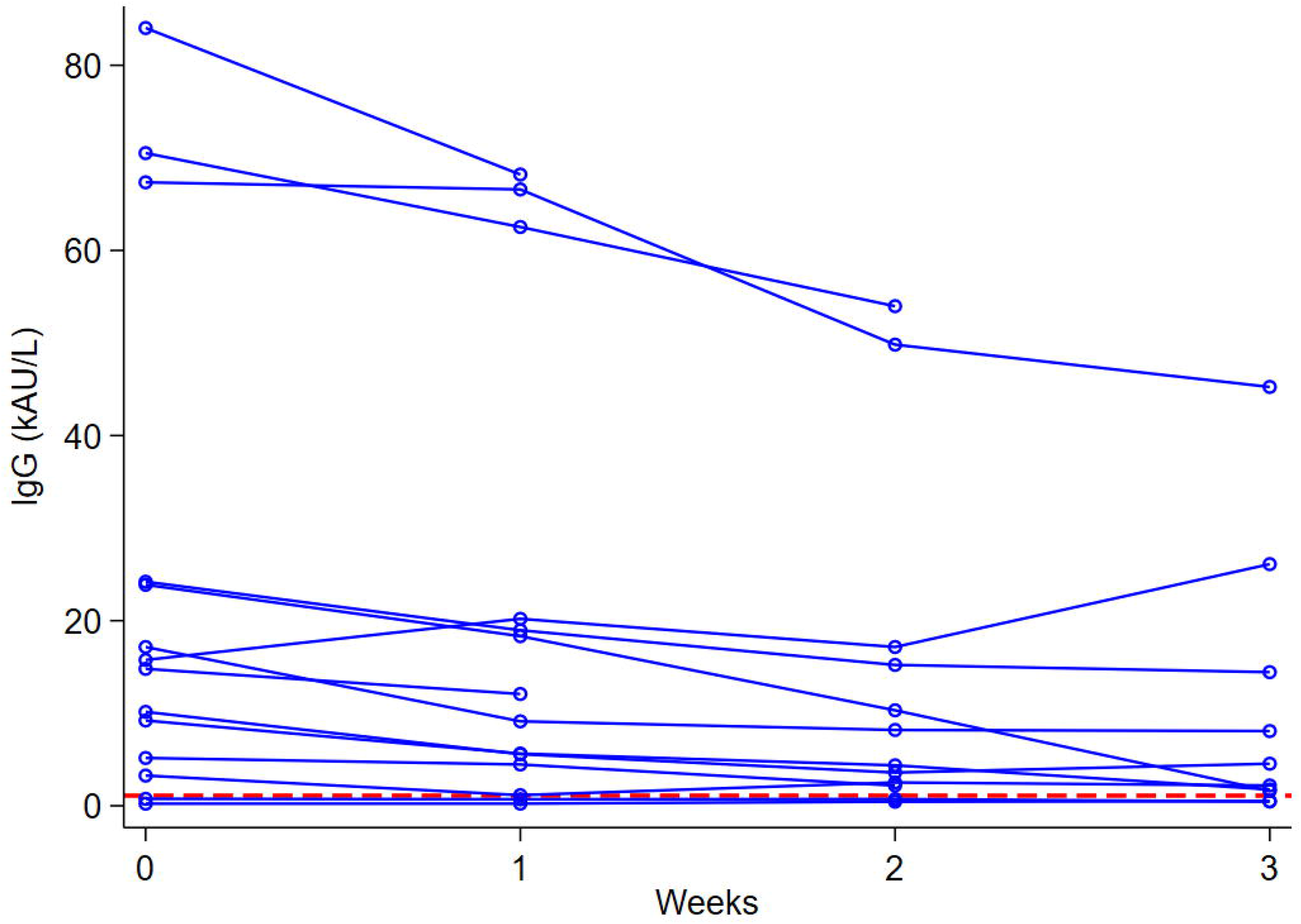

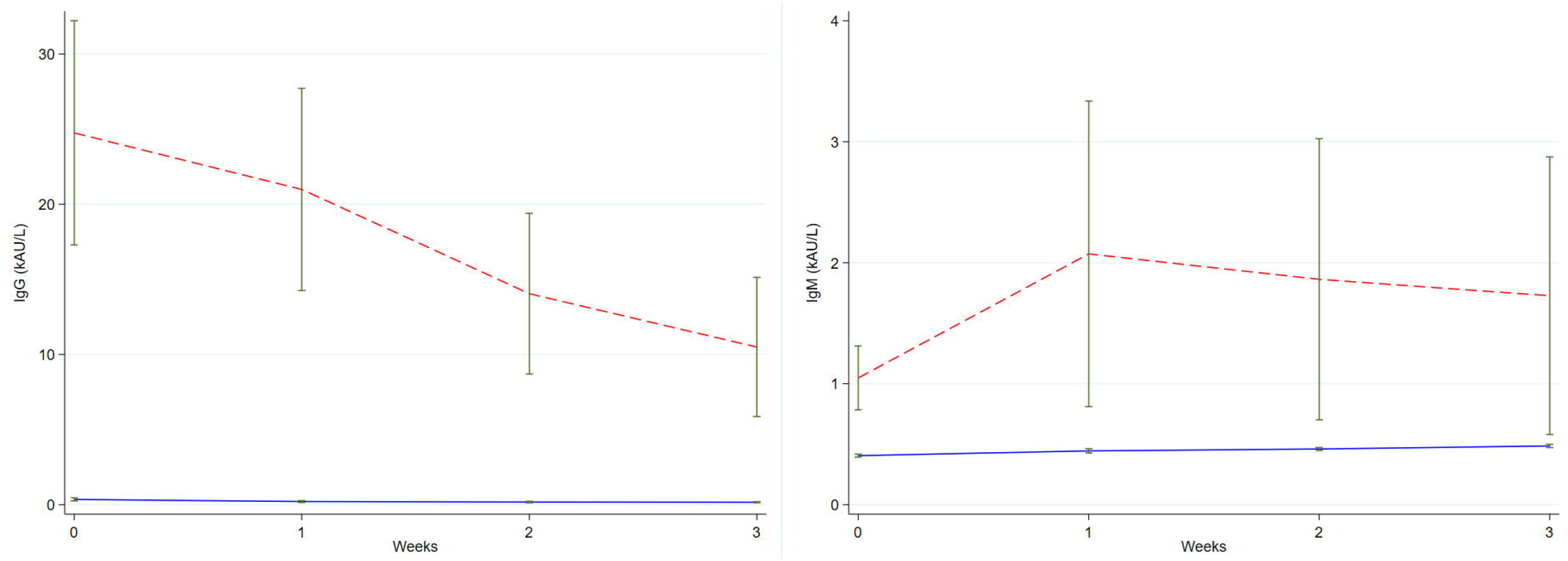

## DISCUSSION

This prospective study reports the results of an epidemiological investigation on *first-line* HCWs who participated with keen interest at PdUH, Italy, during the first wave of the SARS-CoV-2 outbreak in spring 2020. Participants were followed up and tested 4 times for both SARS-CoV-2 using nasopharyngeal swabs and serology every week. Additionally, positive cases were tested again after 5 months. A questionnaire about COVID-19 symptoms and contacts with confirmed cases was administered at every test point.

We found a low incidence of SARS-CoV-2 positivity to nasopharyngeal swabs and serology, 4.07% and 7.85% respectively, in agreement with some reports by other authors as well as a regional sero-survey (6). The low prevalence of SARS-CoV-2 infection in our context could be related to the strict observance of preventive measures against viral transmission among HCWs adopted in our hospital (7). Other studies reported a much higher prevalence of SARS-CoV-2 infections and COVID-19 among HCWs in different settings worldwide (Tab 2).

**Tab 2:** swab-confirmed or anti-SARS-CoV-2antibodies positive HCWs

Tab 2: swab-confirmed or anti-SARS-CoV-2 antibodies positive HCWs
| Authors | Country | COVID-19-confirmed HCWs<br>(pos nasopharyngeal swabs) | % | SARS-CoV-2 Ab pos HCWs | % | Ref |
| --- | --- | --- | --- | --- | --- | --- |
| Moscola et al (40329 HCWs) | USA | 2186/6078<sup>§</sup> | 34.8 | 5523/40329 | 13.7 | 21 |
| Fusco et al | Italy | 2/115 asymptomatic | 1.7 | 2/115 | 1.7 | 11 |
| Lahner et al | Italy | 58/2057 | 2.7 | 8/1084 | 0.7 | 12 |
| Zhao et al (1407 HCWs) | China | 122/152 symptomatic<br>38/908 asymptomatic<sup>#</sup> | 80.3<br>4.2 | 88/908 | 9.7 | 19 |
| Iversen et al | Denmark | np* | / | 1163/28792 | 4 | 22 |
| Porru et al | Italy | 238/5942 symptomatic<br>109/4695 asymptomatic | 4<br>2.3 | np | / | 14 |
| Rivett et al | UK | 31/1032 asymptomatic | 3 | np | / | 13 |
| Durante-Mangoni et al. | Italy | 4/117 | 3.4 | np | / | 23 |
| Stubblefield et al | USA | 3/35 symptomatic | 8.6 | 19/249 | 7.6 | 24 |

\*np = not performed
§ = 93.5% SARS-CoV-2- Ab positive

According to the Italian Health Institute and the Federation of Italian Physicians (Federazione Nazionale degli Ordini dei Medici Chirurghi e degli Odontoiatri), as of 24^th^ November 2020, 64,475 HCWs have been infected with SARS-CoV-2 in Italy, accounting for 0.04% of all cases, and at least 216 physicians have died of COVID-19 (8,9). Lapolla and collaborators reported that these were mainly general practitioners visiting suspected or confirmed cases without adequate PPE (10). In a meta-analysis of data regarding 119,000 COVID-19 patients collected from 11 studies from different countries (China, Italy and the U.S.A.), HCWs counted up to 10.1% of the total positive cases (range: 4.2-17.1) (11). One of the American studies has pointed out a significant rate of HCW SARS-CoV-2 infection out of the hospital, up to 50% (12) and, on average, a reduced HCW COVID-19-related clinical severity and mortality when compared to the general population; this may be due to the younger median age and the absence of relevant comorbidities according to the authors (11).

None of the HCWs included in this study have died of SARS-CoV-2, none have been admitted to Intensive Care Unit, and only two HCWs have been admitted to IDU for pneumonia and discharged without *sequelae*. Among cases, the most common symptoms were fever (OR 12.61, CI95% 1.07-148.2, p = 0.044), nasal congestion (OR 4.72, CI95% 1.21-18.45, p = 0.025), and diarrhoea (OR 10.8, CI95% 1.90-61.6, p = 0.007), while the most common variable related to infection resulted from previous contacts with SARS-CoV-2 individuals (OR 22.2, CI95% 4.42-111.53, p <0.001). The molecular test by nasopharyngeal swab remains the gold standard diagnostic technique for COVID-19, while SARS-CoV-2 immune response detection has proven useful to explore the viral circulation in the general population and the degree of HCW exposure in high risk settings (13, 14, 15). Our serological findings are similar to other reports (Tab 5). The vast majority (eighty-seven percent) of participants positive to SARS-CoV-2 by swab developed antibodies (IgM and /or IgG), as expected.

According to our classification (see M&M), participants in our study were divided in four classes (Tab 3): positive to swab only, (class 1); positive to serology only, (class 2); double positive, (class 3); double negative individuals, (class 4).

**Tab 3:**
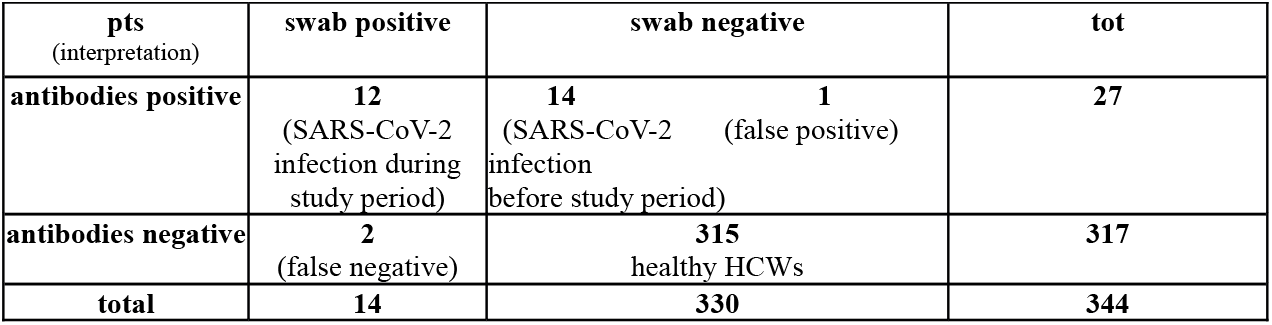
study results and patient outcome classification

**Class 1**: 2 HCWs were positive once to swab, without seroconversion and have to be considered false negatives, in agreement with Fusco et al. (2/115 HCWs; 1,8%) (13).

**Class 2:** 14 HCWs were positive to SARS-CoV-2 antibodies only, with repeated negativity to swabs. In 1 HCW, the positivity to SARS-CoV-2 antibodies only was related to a significant comorbidity (multiple sclerosis) and therefore has been interpreted as an analytic interference and excluded from the study analysis. The remaining 13 cases of this class had an asymptomatic or paucisymptomatic infection, before time zero of our study.

**Class 3:**12 HCWs were positive to both SARS-CoV-2 nasopharyngeal swab and anti SARS-Cov-2 antibodies, and therefore must be considered SARS-CoV-2 acute phase infections during the study period.

**Class 4:** The remaining 315 HCWs, negative to both swab and antibodies, can be considered SARS-Cov-2 free throughout the entire study period.

We found no correlation between gender, professional qualification and place of work with SARS-CoV-2 infection in HCWs, in agreement with other studies (14, 16). The logistic regression analysis showed no significant differences for study variables (place of work, professional qualification, development and type of symptoms, previous contact with confirmed COVID-19 patients, relevant comorbidities and past seasonal flu vaccination) in Abs+ cases, regardless of positive or negative swab. None of Abs+ cases developed a new infection and 89% (24/27) of them had persistent IgG after 5 months. This is in accordance with similar reports (17) and was confirmed by three different Ab tests, whose neutralizing activity is currently under evaluation in our lab (data not shown). As for adaptive immunity, the hypothesis of IgG being a protective immune response to a pathogen goes back more than a century and nevertheless remains even now a fundamental biological principle to establish (18).

The concept behind this and similar studies is not only a safety issue. It also deals with potential benefits and current costs. Testing asymptomatic and symptomatic HCWs is worthwhile to avoid workforce depletion in settings (i.e., Emergency and Infectious Disease Units) that are already at full stretch. It has been reported that in the USA more than 125,000 HCWs were unnecessarily self-isolating due to the lack of specificity of SARS-CoV-2 symptoms. On the other hand, the high number of asymptomatic cases rendersasymptomatic personnel a significantly underestimated potential source of contagion. In 3 independent studies dealing with extensive testing of closed populations (i.e., Vo’ Euganeo, the Diamond Princess cruise ship and the Icelandic population), the number of asymptomatic SARS-CoV-2 positive individuals exceeded 40%, and more than 70% of cases positive to swabs had no symptoms or mild disease (4, 19, 20).

Consistently, other studies reported asymptomatic SARS-CoV-2 infections varying between 1.7% and 4.2%, of all tested HCWs, independently of infection source (i.e., acquired inside the hospital or in the household setting) and accounting for nearly half of all of the positive HCWs (13, 15, 16, 21). Despite all of the fears during the first wave of the SARS-CoV-2 epidemic, evidence is showing an overall reduced risk of COVID-19 for HCWs, highlighting the chance of retrieving effective surveillance protocols from daily experience (1, 15, 16).

A significantly increased risk, up to 3-6 times, of COVID-19 for HCWs compared with the general population was initially reported because of contacts with COVID-19 patients, inadequate use of PPE, management of infective patients and underestimation of viral diffusion on the part of the staff (22, 23). The risk of HCW SARS-CoV-2 infection was significantly increased for staff on duty in non-COVID-19 wards due to inexperience and lack of training (22). In our IDU AT, between February 21th and April 16th 2020, 12,822 SARS-CoV-2 nasopharyngeal swabs were performed on variably symptomatic patients with a daily average of 228 swabs and a rate of swab positivity of 4.3% (544/12.822 pts). 60 HCWs were on duty in this daily service and were regularly tested with anaverage of 6 swabs each (range 4.3-7.1, total number of swabs performed on HCWs 361). No cases of COVID-19 were detected among HCWs during all ofthe IDU AT activity (1).

In conclusion, this study depicts the effect of a strategy to prevent viral spread among HCWs. The measures adopted by our Institution for *first-line* settings were those of pandemics with a high risk of HCW contagion (i.e., comparable to Ebola threat, for whom an educational effort was adopted in 2014) and are part of the “Test, Trace and Isolate” strategy, followed by the Veneto Region where the swabs are available to all contacts of positive cases.

The correct use of PPE, avoiding their re-use, management of confirmed or suspected COVID-19 patients in dedicated areas of the hospital, the virological and serological surveillance of HCWs and the training of staff contributed to the effective control of interpersonal viral spreading. Furthermore, refresh sessions regarding preventive and isolation procedures were planned to avoid staff risk underestimation related to persistent exposure to the virus.

During the first wave of the SARS-CoV-2 spread in spring 2020, we found a low incidence of infected HCWs and could therefore hypothesize the absence of transmission among *first-line* HCWs. Positive HCWs seemed to have been infected outside the hospital, mainly by relatives, as reported by the personal interviews of cases.

Transmission and prevention of infections among HCWs remain issues of global interest, the ongoing spread of SARS-CoV-2 being a current Public Health Emergency of international concern (22-26).

A possible limitation of this study is that it was carried out during the national lock-down of spring 2020 with a limited sample size and single centre design; consequently, our results should be interpreted with caution.

## Data Availability

The copyright remains with the authors. By posting on medRxiv, authors consent to text mining of their work.

## Acknowledgments

We sincerely thank our head nurses, for their endless support and coordination effort: Dott.ssa Suzanne Judet, PS AOUP; Dott.ssa Ilaria Guarnieri, PS OSA; Dott.ssa Lorella Diserò and Dott.ssa Donatella Rampado, IDU AT and Ward.

